# Mapping the association of cerebral small vessel disease, gray matter integrity and cognitive function

**DOI:** 10.1101/2025.02.28.25323102

**Authors:** Marvin Petersen, David Emskötter, Felix L. Nägele, Carola Mayer, Maximilian Schell, Jens Fiehler, Jürgen Gallinat, Simone Kühn, Raphael Twerenbold, Märit Jensen, Götz Thomalla, Bastian Cheng

## Abstract

**INTRODUCTION:** Cerebral small vessel disease (CSVD) is associated with an increased risk of cognitive impairment and dementia. Understanding the brain structural changes associated with CSVD is vital to meet the related health care demands effectively. This study focuses on assessing the integrity of gray matter in relation to CSVD within the general population.

**METHODS & RESULTS:** We examined 2,603 participants (mean age 65 years) from the Hamburg City Health Study, who underwent neuropsychological evaluations and multi-modal neuroimaging. Advanced imaging techniques were used to assess the microstructural and macrostructural integrity of cortical and subcortical gray matter, including the hippocampus. Our findings indicate that the extent of CSVD is associated with abnormalities in gray matter diffusivity, myelin content, and morphology in specific brain regions including the anterior cingulate, insular and temporal cortices, caudate, putamen, pallidum, and hippocampus. Crucially, CSVD-related gray matter abnormalities were linked to cross-domain cognitive performance, represented by the first principal component of multi-domain cognitive test scores.

**DISCUSSION:** Complementing previous research that focuses on CSVD and white matter changes, our study highlights abnormal gray matter integrity as a possible link between small vessel pathology and cognitive disorders. The insights gained can guide diagnostic and therapeutic strategies, supporting the advancement of interventions tailored to mitigate the impact of CSVD.

## Introduction

Cerebral small vessel disease (CSVD) is a relevant contributor to cognitive decline, Alzheimer’s and vascular dementia in ageing populations.^1^ Given the current lack of therapeutic interventions, the identification of disease biomarkers is a key priority in CSVD research for converting advances in pathomechanistic understanding into effective prevention and treatment approaches.^2–4^ Brain magnetic resonance imaging (MRI) is at the forefront of these efforts, enabling the identification of structural brain alterations in all stages of CSVD. Established imaging markers range from altered diffusion measures at the microscopic level to white matter hyperintensities of presumed vascular origin (WMH) as well as enlarged perivascular spaces (PVS), observable in standard anatomical imaging sequences.^5,6^

Despite this progress, considerable gaps in understanding persist: the majority of current imaging analyses in patients with CSVD investigate abnormalities in the cerebral white matter and their clinical consequences.^7,8^ Despite the known impact of CSVD on gray matter integrity,^5^ there is a notable lack of well-powered neuroimaging studies utilizing a more diverse array of imaging markers capturing gray matter structural properties in CSVD.^9–11^ Furthermore, most analyses focus on global brain morphology or predefined regions of interest, rather than examining the full range of gray matter areas, including the cortex, basal ganglia, and hippocampus.^12^ As a result, the relationship of CSVD imaging markers, regional gray matter micro- and macrostructure, and cognition, specifically in early, preclinical stages of CSVD, remains unclear.

We, therefore, argue for holistic approaches that unify large-scale, behavioral as well as imaging data and advanced neuroimaging techniques to comprehensively characterize the pathomechanistic correlates of CSVD.^13,14^ We address this research need by utilizing data from participants of the Hamburg City Health Study (HCHS), a large-scale epidemiological study with extensive clinical and imaging phenotyping.^15^ Specifically, we integrate established CSVD imaging markers and micro- and macrostructural profiling of both the cortex and subcortex based on diffusion imaging, myelin mapping and morphometry with epidemiological data and neuropsychological assessments across multiple cognitive domains. Through this approach, our research aims to advance the understanding of the neural underpinnings of CSVD addressing two hypotheses: (1) An increasing CSVD burden (operationalized by current standards in neuroimaging^5,6^) is linked to regionally specific differences in gray matter micro- and macrostructure; (2) CSVD-related brain structural differences are associated with cognitive function.

## Materials and methods

The methodological approach of this study is illustrated in *figure 1*.

**Figure 1.**
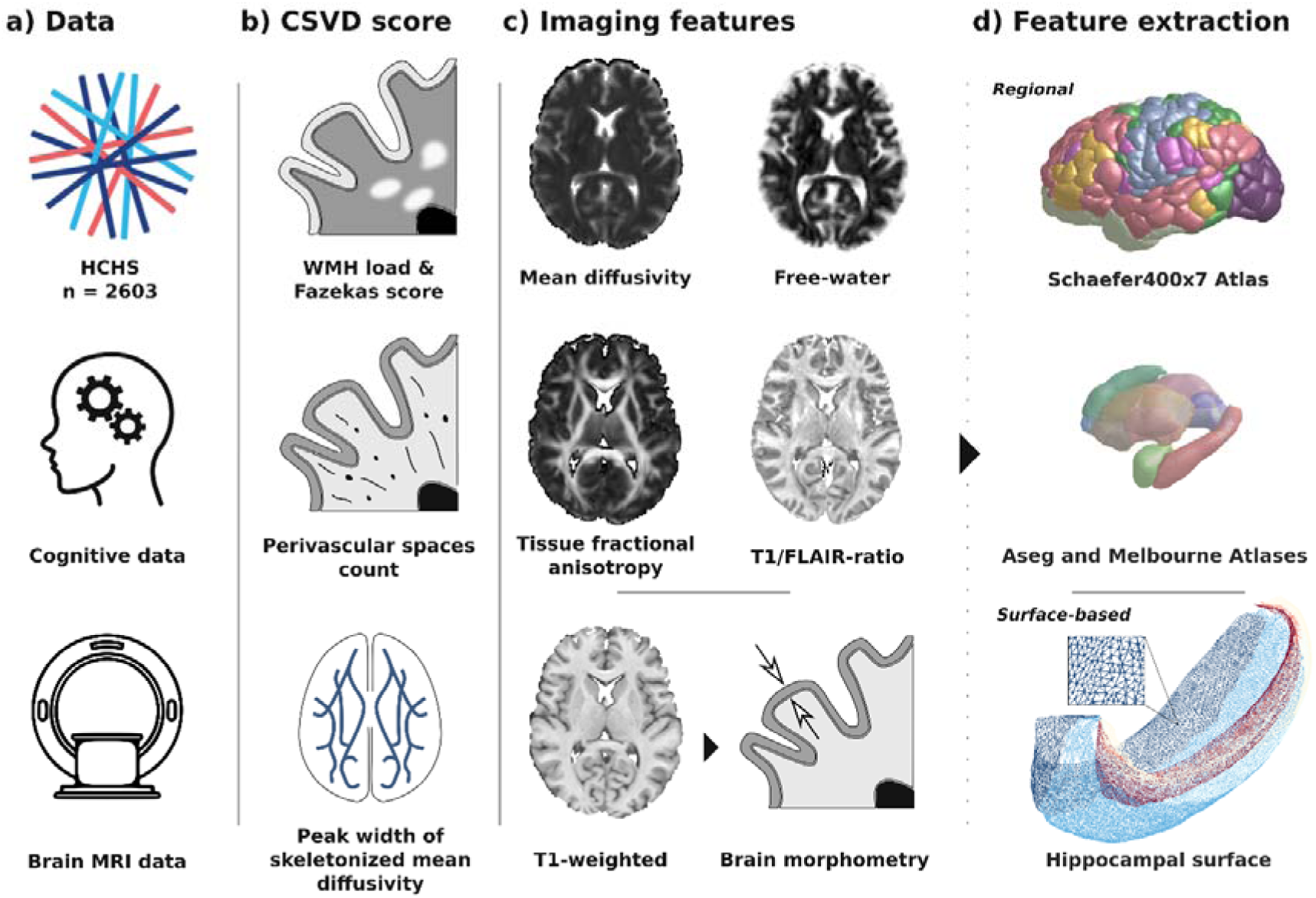
Methodology. a) The analysis was based on cognitive and brain MRI data of the Hamburg City Health Study. b) A CSVD score was computed by averaging the z-scored WMH load, Fazekas score, perivascular spaces count and PSMD. c) Imaging features of brain micro- and macrostructure were derived based on T1-weighted, FLAIR and diffusion-weighted images. Brain morphometry was performed on T1-weighted images to obtain cortical thickness and subcortical volume measures. The T1/FLAIR-ratio was computed as a surrogate of myelin content. Diffusion tensor imaging and free-water imaging were performed to compute indices of gray matter microstructure: mean diffusivity, free-water, and tissue fractional anisotropy. d) For cortical and subcortical areas, feature extraction was performed aggregating imaging features in regions defined by the Schaefer400×7 Atlas, Aseg Atlas and Melbourne Subcortex Atlas.^26,34,41^ For the hippocampus, feature extraction was performed on the vertex-level based on hippocampal surface reconstruction with HippUnfold.^35^ *Abbreviations*: CSVD = cerebral small vessel disease, FLAIR = Fluid-attenuated inversion recovery, HCHS = Hamburg City Health Study, MRI = magnetic resonance imaging, PSMD = peak width of skeletonized mean diffusivity, WMH = white matter hyperintensities of presumed vascular origin.

### Study population

We examined cross-sectional clinical and imaging data of 2631 participants from the HCHS (*figure 1a*).^15^ Individuals with neurological diagnoses based on neuroradiological assessments (e.g., tumors, ischemic stroke) were excluded. The HCHS is a prospective, population-based, single-center cohort study in Hamburg, Germany, investigating adults aged 45-75 to enhance the detection of major chronic disease risks through extensive clinical and imaging phenotyping.^15^ The HCHS was approved by the local ethics committee of the Landesärztekammer Hamburg (State of Hamburg Chamber of Medical Practitioners, PV5131). Good Clinical Practice (GCP), Good Epidemiological Practice (GEP) and the Declaration of Helsinki were the ethical guidelines that governed the conduct of the HCHS.^16^ Written informed consent was obtained from all participants investigated in this work.

### Cognitive assessments

Cognitive functions were assessed via the Animal Naming Test, Multiple Choice Vocabulary Intelligence Test B, Trail Making Test A and B and Word List Recall Test of the extended version of the Consortium to Establish a Registry for Alzheimer’s Disease Neuropsychological Assessment Battery (CERAD-NP/Plus), as well as the Clock Drawing Test.^17,18^ All tests were administered by a trained study nurse. To ensure that a higher value corresponds with higher cognitive performance across all tests, we inverted the Trail Making Test A and B results.

### MRI acquisition and preprocessing

MR imaging was conducted using a single 3-Tesla Siemens Skyra. The scanning protocols follow those outlined in previous studies. For 3D T1-weighted anatomical images, a rapid acquisition gradient-echo sequence (MPRAGE) was used with the following sequence specifics: repetition time (TR) = 2500 ms, echo time (TE) = 2.12 ms, 256 axial slices, slice thickness (ST) = 0.94 mm, in-plane resolution (IPR) = 0.83 × 0.83 mm. 3D T2-weighted fluid-attenuated inversion recovery (FLAIR) images with TR = 4700 ms, TE = 392 ms, 192 axial slices, ST = 0.9 mm, and IPR = 0.75 × 0.75 mm. For single-shell diffusion-weighted imaging (DWI) 75 axial slices were acquired with gradients (b = 1000 s/mm^2^) applied along 64 noncollinear directions with the following sequence parameters: TR = 8500 ms, TE = 75 ms, ST = 2 mm, IPR = 2 × 2 mm, anterior–posterior phase-encoding direction, 1 b0 volume.

MRI data were processed employing preconfigured containerized pipelines as described previously.^14^ The full documentation can be found on GitHub (https://github.com/csi-hamburg/CSIframe/wiki). Diffusion- and T1-weighted images were preprocessed employing QSIprep (v. 0.14.2).^19^ DWI preprocessing included bias field correction, skull-stripping, denoising, deringing, as well as correction of B1 field inhomogeneity, head motion, eddy currents, and susceptibility distortions.^19–23^ T1-weighted and FLAIR images underwent bias field correction and skull-stripping.^23^

### Computation of CSVD score

Following previous recommendations we computed a summary CSVD score from four imaging markers representing CSVD burden: WMH load, Fazekas score, perivascular spaces count and peak width of skeletonized mean diffusivity (*figure 1b*).^6^

WMH were segmented using a fully automated pipeline as described previously.^14^ In brief, FSL’s Brain Intensity AbNormality Classification Algorithm (BIANCA) was employed with LOCally Adaptive Threshold Estimation (LOCATE) on preprocessed FLAIR images and T1-weighted images.^24,25^ After applying both algorithms, the segmentations were refined using Freesurfer v.6.0.1 parcellations to exclude non-white matter regions.^26^ WMH load was calculated by normalization of WMH volume for intracranial volume.^26^

Fazekas scoring was performed on FLAIR images by trained neuroradiologists following standard procedures.^27^ Briefly, the Fazekas scale grades WMH as 0 (absence of WMH), 1 (punctate WMH), 2 (early confluent WMH), or 3 (WMH in large confluent areas). Periventricular and deep white matter are scored separately. A Fazekas sumscore was computed for each subject by summing deep and periventricular subscores.

PVS were automatically segmented based on T1-weighted images applying a validated method based on Frangi filtering for vessel identification.^28–30^ Applied Frangi filter parameters were α = 0.5, β = 0.5 and c = 500 following previous recommendations.^30^ Resulting vessel-shape likelihood maps were locally thresholded to obtain PVS segmentations. Applied local thresholds for the basal ganglia, centrum semiovale and midbrain were previously optimized based on visual inspection of a randomly chosen subsample of n = 100 participants. For quality assessment, PVS masks were visually inspected and a subsample of 100 participants – 25 randomly chosen participants per PVS count quartile – was assessed with a visual rating scale by a trained expert (B.C.). Scale items are 0 (no PVS), 1 (mild; 1–10 PVS), 2 (moderate; 11–20 PVS), 3 (frequent; 21–40 PVS) or 4 (severe; >40 PVS).^31^ The correlation between PVS counts and PVS visual rating scores was r = 0.62.

The peak width of skeletonized mean diffusivity (PSMD) was calculated based on standard procedures and adapted for our purposes by using non-linear registration with the Advanced Normalization Tools (ANTs) SyN registration.^32,33^ PSMD was calculated as the difference between the 95th and 5th percentile of MD values on the white matter skeleton in standard (MNI) space.

Missing values for the CSVD measures were mean imputed. The CSVD score was derived by averaging the z-scores of WMH load, Fazekas score, PVS count, and PSMD. The counts of microbleeds and recent small subcortical infarcts were not considered for the CSVD score computation given a relevant proportion of missing data and occurrence in few subjects. A correlation matrix of the CSVD score and its subcomponents is shown in *supplementary figure S2*.

### Imaging features of gray matter micro- and macrostructure

Five measures representing different aspects of gray matter micro- and macrostructure were computed (*figure 1c*): thickness & volume representing brain morphology; T1/FLAIR-ratio representing myelination; mean diffusivity (MD) representing bulk water diffusivity; free-water representing volume of the extracellular compartment; and tissue fractional anisotropy representing directionality of water diffusion attributable to the tissue compartment. All gray matter measures were extracted for the following regions: cortical regions defined by the Schaefer400×7 Atlas;^34^ subcortical regions including the caudate, nucleus accumbens, amygdala, pallidum, putamen, and thalamus; and vertices of the hippocampal midthickness surface derived via HippUnfold.^35^

#### Intensity-based measures

Diffusion imaging. Diffusion imaging features were derived from preprocessed diffusion-weighted images. Diffusion-tensor imaging was performed to compute voxel-wise maps of mean diffusivity. For this, diffusion tensors were modelled based on preprocessed DWI using a least-squares fit.^36,37^ Furthermore, we conducted free-water imaging, employing a dual-tensor model that delineates the isotropic extracellular compartment and the cellular compartment characterized by hindered or restricted diffusion. Based on a regularized non-linear fitting process, free-water and tissue fractional anisotropy (FA_T_) were calculated.^38^

T1/FLAIR-ratio. The T1/FLAIR-ratio was computed following previous procedures.^39,40^ The T1-weighted and FLAIR images were non-linearly registered to the MNI152NLin2009cAsym standard space.^33^ The normalized T1-weighted image was divided by the FLAIR image.

The intensity-based measures were further mapped from the voxel level to cortical and subcortical regions as well as hippocampal surface vertices. Cortical and subcortical measures were obtained by averaging within regions of interest defined by the Schaefer400×7 Atlas and Melbourne Subcortex Atlas.^34,41^ As the Melbourne Subcortex Atlas divides the thalamus in an anterior and posterior part, these regions were subsequently averaged to obtain a single value for the thalamus per measure per subject. As a result, intensity-based measures were derived for the same subcortical structures as volumes based on the Aseg atlas. Furthermore, voxel measures were mapped to vertices of the hippocampal and dentate gyrus surfaces in MNI152NLin2009cAsym (0p5mm, https://github.com/khanlab/hippunfold-templateflow) using the Connectome Workbench (v. 1.5.0).^42^ For the hippocampal surface, ribbon-constrained volume to surface mapping was employed using the inner, outer and midthickness surfaces.^43^ For the dentate gyrus surface trilinear interpolation volume to surface mapping was performed.^43^ Subsequently, surface data were smoothed with a gaussian kernel (σ = 2).

#### Morphometry

Thickness and volume. FreeSurfer (v. 6.0.1) was used to derive regional measures of cortical thickness and subcortical volumes.^26,44^ After surface reconstruction, cortical thickness was measured as the distance between the white matter and pial surface.^44^ Vertex-wise hippocampal thickness was measured in native space using HippUnfold (v. 1.2.0).^35^ HippUnfold processing includes preprocessing and cropping around the hippocampi, deep learning-based tissue class segmentation, unfolding, computation of a mapping between native Cartesian space and unfolded space, and transformation of standardized unfolded surfaces to native space.^45,46^ Surface reconstructions resulting from HippUnfold include the inner, midthickness and outer surface as well as the dentate gyrus surface. Hippocampal thickness was computed using Connectome Workbench as the distance between inner and outer surface.^43^ HippUnfold does not perform thickness measurements for the dentate gyrus.

### Quality assessment

Quality assessment included visual inspection of raw images and processed images based on quantitative outliers defined as measures exceeding 2 standard deviations from the mean of quality measures computed via QSIprep and mriqc.^19,47^ In addition, all HippUnfold outputs were visually inspected. Images with insufficient quality were excluded. A flowchart detailing the sample selection procedure is shown in *supplementary figure S1*.

### Statistical analysis

All statistical analyses were conducted in python 3.9.1 leveraging Pingouin (v. 0.5.4) as well as FSL’s Permutation Analysis of Linear Models (PALM) based on Matlab v.2021b.^48^ Statistical tests were two-sided, with a *P*<0.05 as significance threshold. *P-*values were adjusted via family wise error correction for multiple comparisons.

#### Regional association of CSVD score and imaging features

To investigate the association of CSVD burden and gray matter integrity, we conducted permutation-based testing for linear associations between the CSVD score and regional features of gray matter micro- and macrostructure in a general linear model. Statistical tests were two-sided (n_permutation_ = 5000), with a p < 0.05 as the significance threshold. To account for multiple comparisons, p*-*values were adjusted for family-wise error. General linear models were adjusted for age, sex and education. To obtain standardized β-coefficients, input variables were z-scored beforehand. As a result, β-coefficients and p-values were obtained for each cortical and subcortical region as well as each hippocampal vertex indicating the strength and significance of the CSVD score’s linear association with respective imaging features for each ROI.

#### Associations between imaging features and cognitive functioning

Cognitive test scores were dimensionality reduced via a principal component analysis. The first principal component explaining the highest amount of variance was defined as a measure of *general cognitive ability* (*g*) following previous procedures.^49^ A correlation matrix of g and the cognitive test scores is shown in *supplementary figure S3*.

To obtain a single aggregate measure per subject per feature, imaging features were averaged across cortical and subcortical regions with a significant association of the respective imaging index and the CSVD score. This means that any region demonstrating a significant correlation between the CSVD score and an imaging feature was included in the aggregation process for that specific feature. Prior to this, imaging features were averaged across hippocampal vertices to produce a single measurement for each subject for both the left and right hippocampus. Aggregate imaging scores were associated with *general cognitive ability* (*g*) using multiple linear regression adjusted for age, sex and education.

### Data availability

HCHS data can be obtained by qualified researcher on reasonable request to the study’s steering committee.

## Results

### Sample characteristics

The final analysis sample included 2,603 individuals (44% female, median [IQR] age 65 [14] years; for details see *table 1*). For a flow chart on the sample selection procedure refer to supplementary *figure S1*.

**Table 1.** Sample characteristics.

| Table 1. Sample characteristics |  |
| --- | --- |
| Metric | Stat<sup>a</sup> |
| Age, years | 65 [14] (2,603) |
| Female, n (% female) | 1,144 (44) |
| Education, ISCED | 2 [1] (2,603) |
| **CSVD markers** |  |
| WMH load, % | 0.001 [0.001] (2,603) |
| Fazekas scores | 1 [2] (2,603) |
| Perivascular spaces count, n | 66 [61] (2,603) |
| Peak width of skeletonized mean diffusivity, 10<sup>-3</sup> mm<sup>2</sup>/s | 0.23 [0.04] (2,603) |
| **Global imaging features** |  |
| Mean diffusivity, 10<sup>-2</sup> mm<sup>2</sup>/s | 0.12 [0.01] (2,398) |
| Free-water | 0.51 [0.07] (2,398) |
| Tissue fractional anisotropy | 0.19 [0.01] (2,398) |
| T1/FLAIR-ratio | 5.73 [2.54] (2,554) |
| Mean cortical thickness, mm | 2.63 [0.13] (2,550) |
| Mean subcortical volume, ml | 1296.21 [174.35] (2,550) |
| **Cognitive test scores** |  |
| Animal Naming Test | 24 [9] (2,444) |
| Clock Drawing Test | 7 [7] (2,501) |
| Multiple Choice Vocabulary Intelligence Test | 32 [4] (2,044) |
| Trail Making Test A, sec | 37 [17] (2,314) |
| Trail Making Test B, sec | 82 [42] (2,288) |
| Word List Recall | 8 [2] (2,369) |
| Abbreviations: ISCED = International Standard Classification of Education, mm = millimeters, sec = seconds |  |
| <sup>a</sup>Presented as median [IQR] (N) |  |

a) Data

### Regional association of CSVD score and imaging features

To investigate gray matter integrity differences in relation to CSVD burden, we performed a region-level analysis of the relationship between the CSVD score and imaging features of gray matter micro- and macrostructure (*figure 2)*. Higher CSVD scores were related to regional differences in brain structural features – i.e., higher mean diffusivity and free-water in the lateral prefrontal, cingulate, insular, temporal and occipital cortices as well as all subcortical structures; lower mean diffusivity bilaterally in the superior frontal and parietal cortices (*figure 2a* and *b*); lower tissue fractional anisotropy bilaterally in the anterior cingulate and insular cortices, caudate, thalamus, the cornu ammonis and dentate gyrus of the hippocampus and the left amygdala; higher tissue fractional anisotropy bilaterally in the putamen (*figure 2c*); lower T1/FLAIR-ratio bilaterally in the temporal cortex and hippocampus as well as the right amygdala (*figure 2d*); lower cortical thickness bilaterally in the anterior cingulate, insular and temporal cortices as well as lower volume in the nucleus accumbens; higher volumes in the caudate, putamen and pallidum as well as higher thickness in the subiculum and cornu ammonis of the hippocampus (*figure 2e*).

**Figure 2.**
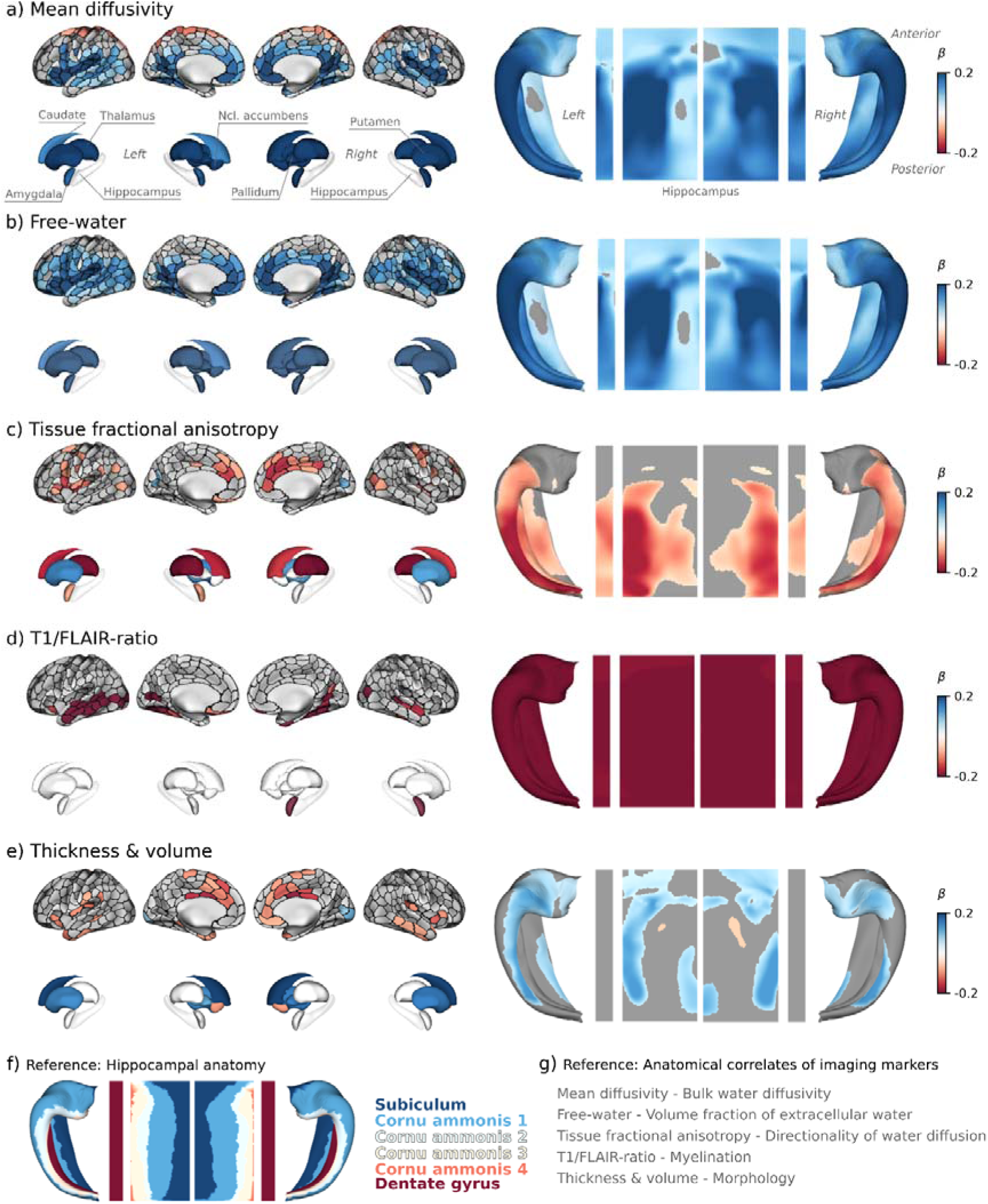
Regional associations of CSVD score and imaging features. Anatomical plots display the regional relationship between the CSVD scores and imaging features. Regional associations were investigated in cortical ROIs defined by the Schaefer400 Atlas, subcortical ROIs defined by the Melbourne Subcortex Atlas and vertices on the hippocampal surface. ROIs / vertices in which the CSVD score were significantly associated with regional imaging features after family-wise error-correction are highlighted by colors encoding standardized β-coefficients from general linear models: a negative β (red) denotes that a higher CSVD score, i.e., higher cumulative burden of CSVD markers, is associated to a lower regional imaging index; a positive β (blue) indicates that a higher CSVD score is linked to a higher regional imaging index. To facilitate effect comparisons between the different imaging features, colormaps were thresholded at β values of −0.2 and 0.2. Each facet displays cortical ROIs in the top row, subcortical ROIs in the middle row, and hippocampal surfaces (folded and unfolded) in the bottom row. Statistics were not performed for the hippocampus on the ROI level, only on the hippocampal surface. Therefore, the hippocampal ROI in the middle row is faded out. Facets a) – e) correspond with a different imaging index: a) Mean diffusivity, b) free-water, c) tissue fractional anisotropy, d) T1/FLAIR-ratio, e) cortical thickness, subcortical volume and hippocampal thickness. Facet f) and g) illustrate hippocampal reference anatomy and list anatomical correlates of the investigated imaging features.

### Cognition analysis of imaging indices

The approach for the analysis of cognitive data is illustrated in *figure 3a*. To obtain a measure of general cognitive ability, cognitive test scores were subjected to a principal component analysis. The first principal component (PC1) explained 35.0% of variance in cognitive data (*figure 3b*) and loadings indicated that all cognitive test scores contributed to this variable (*figure 3c*) confirming it as a measure of general cognitive ability. In multiple linear regression models adjusted for age, sex and education, a higher PC1 score – i.e., lower general cognitive ability – was significantly associated with higher mean diffusivity (std. β = 0.140, *P_FWE_* < 0.001), higher free-water (std. β = 0.183, *P_FWE_* < 0.001), and lower tissue fractional anisotropy (std. β = −0.097, *P_FWE_* < 0.001), lower T1/FLAIR-ratio (std. β = −0.090, *P_FWE_* < 0.001), lower thickness & volume (std. β = −0.051, *P_FWE_* = 0.036) (*figure 3d*). For regression plots displaying the association between aggregate imaging indices and individual cognitive test scores results see *supplementary figure S4*.

**Figure 3.**
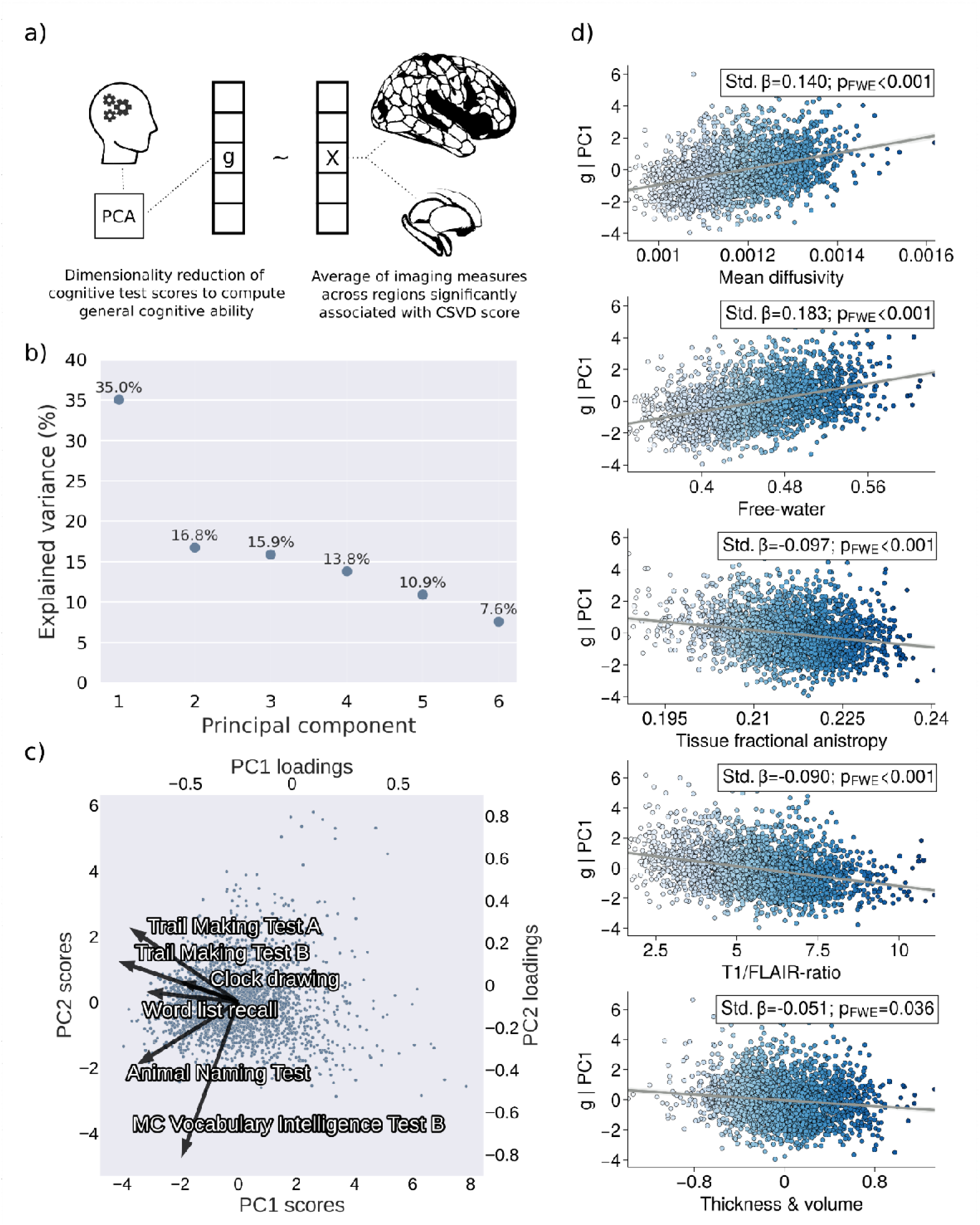
Clinical analysis. a) Approach. Imaging features were averaged across cortical and subcortical regions with a significant association of the respective imaging index and the CSVD score. Cognitive test scores were dimensionality reduced via a principal component analysis. The first principal component explaining the highest amount of variance was defined as a measure of general cognitive ability (g). Averaged imaging scores were associated with g using multiple linear regression adjusted for age, sex and education. b) Scree plot indicating explained variance for each principal component. c) Biplot indicating PCA loadings for each cognitive test score alongside subject-level scores. d) Regression plots displaying the association between aggregate imaging features and g. *Abbreviations*: g = measure of general cognitive ability corresponding with principal component 1 of the PCA of cognitive test scores, MC = multiple choice, PC = principal component, PCA – principal component analysis, p_FWE_ = family wise error-corrected p-value, std. β – standardized β coefficient derived from multiple linear regression.

## Discussion

In a large population-based sample we conducted an in-depth investigation of the association between CSVD burden, gray matter integrity and cognitive function. We report on two main findings: (1) a higher CSVD burden – operationalized by current neuroimaging standards – was linked to regionally specific differences in gray matter micro- and macrostructure; (2) CSVD-related gray matter abnormalities were associated to cognitive performance. Overall, our findings identify a complex profile of structural brain changes, offering novel insights into the relationship between small vessel pathology and brain health.

### CSVD is linked to regionally specific differences in gray matter micro- and macrostructure

CSVD compromises brain health through intricate, interconnected effects on the cerebral parenchyma. To characterize CSVD-related abnormalities in gray matter integrity we investigated advanced neuroimaging markers of tissue micro- and macrostructure.

Our analysis showed that a higher CSVD burden, represented by a higher CSVD score, was associated with higher gray matter mean diffusivity reflecting higher bulk water movement, higher extracellular free-water, and lower fractional anisotropy, which suggests a decrease in the directional movement of water molecules within brain tissue. These findings confirm the sensitivity of diffusion measures to CSVD-related pathology demonstrated in the white matter and extend these observations to gray matter regions.^7,8,50^ Differences in mean diffusivity and free-water were widespread spanning the anterior cingulate, insular, temporal and occipital cortices as well as all investigated subcortical areas. We hypothesize that higher overall water diffusivity and extracellular free-water in CSVD are attributed to CSVD-related blood-brain barrier disruptions and inflammation-induced osmotic shifts of water from blood to the extracellular space.^51^ Of note, the pattern of CSVD-related diffusivity changes, specifically the involvement of temporal and cingulate areas, coincided relevantly with effect maps recently reported for Alzheimer’s disease and atrial fibrillation, consistent with the increasingly recognized pathomechanistic overlap across these conditions.^52,53^ Yet, previous work shows that in the white matter CSVD determines abnormalities in diffusion MRI stronger than Alzheimer’s pathology which might indicate that CSVD pathophysiology is the most parsimonious explanation to the observed shifts in tissue diffusivity.^54^

The differences in tissue fractional anisotropy were primarily observed in more spatially limited areas, specifically the cingulate and insular cortices, the caudate, putamen, thalamus, amygdala, and hippocampus. As histopathological studies have indicated that gray matter fractional anisotropy corresponds with non-myelinated neurite integrity,^55^ our findings could suggest that the highlighted regions exhibit cellular pathology beyond volume shifts in the extracellular compartment, i.e., cellular disruption and abnormalities in neurite architecture. Collectively, regional abnormalities of microstructural imaging markers showed pronounced symmetry, which we interpret as confirmatory of vascular contributions, given the symmetric distribution of vascular supply.

Lastly, we identified a reduced T1/FLAIR-ratio bilaterally in the temporal cortex and hippocampus, indicating decreased myelin content in these regions.^40^ Although prior studies demonstrated an association of CSVD with temporal lobe atrophy,^56–58^ investigations into gray matter myelin content were not conducted before. Since the temporal lobe is a critical area affected by neurodegeneration in Alzheimer’s disease,^59^ our findings may represent an interaction pathway between Alzheimer’s and vascular pathology.^60^ Supporting this, transgenic mice with Alzheimer’s disease show altered expression of myelin markers in the hippocampus and entorhinal cortex preceding cognitive decline.^61,62^ Additionally, a neuroimaging study using myelin mapping has found reduced myelin content in the medial temporal lobes of Alzheimer’s patients.^63^ Importantly, animal research suggests that demyelination might precede and facilitate amyloid deposition in Alzheimer’s disease: myelin dysfunction aggravates the accumulation of amyloid β-producing elements and the increase in amyloid precursor protein cleavage.^64^ Therefore, CSVD-induced demyelination in vital temporal lobe structures could accelerate dementia progression. Maintaining myelin integrity by preventing CSVD may thus be an effective approach to delay dementia onset.

### Higher CSVD burden links to abnormal brain morphology

Turning to morphometric measures, we found altered cortical thickness, subcortical volumes and hippocampal thickness associated with CSVD burden. Regionally, we found CSVD-related lower cortical thickness to be localized bilaterally to the anterior cingulate, insular and temporal cortices indicative of neurodegenerative effects. CSVD may lead to a reduction in cortical thickness by driving multiple vascular and inflammatory mechanisms, that hinder proper blood flow and oxygenation, thereby accelerating neuronal tissue loss.^2^ In addition, altered cortical morphology might be a result of WMH-related disconnectivity.^65^ Importantly, lower cortical thickness and lower tissue fractional anisotropy coincided within the anterior cingulate and insula which further underscores that these regions may exhibit more severe CSVD-related tissue damage. These findings support previous evidence indicating cortical atrophy in CSVD.^10,11^

In contrast, subcortical structures largely showed higher volume and hippocampal thickness accompanying higher CSVD burden. Although speculative, these disparities might be a result of varying local stages of tissue changes all tethered to the shared disease mechanism of small vessel injury. This interpretation finds support in previous work assessing cortical thickness in multiple stages of neurodegeneration due to Alzheimer’s dementia.^66^ The corresponding findings indicate a biphasic trajectory of cortical thickness changes in Alzheimer’s disease: in early preclinical disease stages cortical thickening was observed while in later stages cortical thinning was evident. These findings may also apply to CSVD, where subcortical areas might show subtler pathology primarily evident at the microstructural level without clear morphological manifestations. Our observations of widespread microstructural differences in the cortex and subcortex align with this, potentially acting as precursors to more pronounced neurodegenerative processes. Mechanistically, higher amounts of extracellular free-water might correspond with subtle edema leading to a measurable increase of thickness and volume. Thus, the regions showing microstructural deviations in our current cross-sectional analysis might manifest macrostructural decline as the disease progresses. Longitudinal neuroimaging studies are required to further substantiate these hypotheses in CSVD.

### CSVD-related gray matter abnormalities are linked to cognitive function

There are many studies demonstrating interactions between CSVD and cognitive disorders, spanning mild cognitive impairment to Alzheimer’s and vascular dementia.^67^ To characterize the contribution of CSVD-related differences in gray matter integrity to variance in cognitive performance, we performed post-hoc multiple linear regression analyses relating aggregated imaging features and general cognitive ability. Prominently, all aggregate imaging features were significantly associated with general cognitive ability represented by *g* as construct that represents cross-domain cognitive function. Worse general cognitive ability related to higher gray matter mean diffusivity and free-water as well as lower tissue fractional anisotropy, T1/FLAIR-ratio, thickness and volume. On the level of individual cognitive test scores, shifts in gray matter integrity were associated with lower performance in executive function (Trail Making Test B), information processing speed (Trail Making Test A), reasoning (Multiple Choice Vocabulary Intelligence Test), and verbal fluency (Animal Naming Test) (*supplementary figure S4*).

These findings shed light on the complex interactions between CSVD, neuroanatomy and cognitive impairment, suggesting potential clinical applications. Although the effectiveness of CSVD treatments in preventing cognitive consequences remains to be conclusively proven, promising results from risk factor adjustments, especially in blood pressure management,^68,69^ suggest improved cognitive outcomes. Future clinical trials on CSVD require reliable biomarkers to accurately identify vascular contributions to cognitive decline and pinpoint at-risk individuals.^3,4^ Integrating data on gray matter micro- and macrostructure with established CSVD indicators could enhance our understanding of the underlying mechanisms, enabling better identification of those most likely to benefit from targeted interventions.

### Strengths and limitations

The strength of our study is the integration of large-scale clinical and multimodal MRI data with advanced analysis techniques, offering a detailed view of the association of CSVD with brain health in the normal population. However, our cross-sectional design may limit deeper insights into CSVD and cognition dynamics that only a longitudinal approach might unveil. Our definition of CSVD burden is limited to markers applicable to a population-based context. Nonetheless, considering further CSVD markers in patient populations including recent small subcortical infarcts, cortical microinfarcts and cerebral microbleeds may provide complementary insights. Lastly, the focus on a single population of mainly European ancestry may limit the generalizability of our results to other populations.

## Conclusion

Our comprehensive investigation of a large population-based cohort revealed links between CSVD, abnormalities of gray matter integrity, and cognitive function. Our research emphasizes that small vessel pathology appears to impact micro- and macrostructural integrity of cortical and subcortical areas contributing to cognitive deficits. As this research field progresses, harnessing neuroimaging could pave the way for individualized diagnostics and therapeutic approaches in CSVD.

## Data Availability

HCHS data can be obtained by qualified researcher on reasonable request to the studys steering committee.

## Acknowledgments

The authors would like to acknowledge all participants, all cooperation partners, patrons and the Deanery from the University Medical Center Hamburg-Eppendorf for supporting the HCHS. Special thanks are due to the staff at the Population Health Research Department for conducting the study. The publication of its results has been approved by the Steering Board of the HCHS.

## Author Contributions

We describe contributions to the paper using the CRediT contributor role taxonomy. M.P.: Conceptualization, Data Curation, Formal analysis, Investigation, Methodology, Project administration, Resources, Software, Visualization, Writing—original draft, Writing—review & editing; D.E.: Data curation, Software, Writing—review & editing; F.L.N.: Data curation, Software, Writing—review & editing; C.M.: Data curation, Software, Writing—review & editing; M.S.: Data curation, Software, Writing—review & editing; J.G.: Writing—review & editing; S.K.: Writing—review & editing; R.T.: Resources, Writing—review & editing; M.J.: Writing—review & editing; G.T.: Funding acquisition, Supervision, Writing—review & editing; B.C.: Data curation; Conceptualization, Funding acquisition, Supervision, Writing—review & editing.

