## Supplementary materials for "Mapping the association of cerebral small vessel disease, gray matter integrity and cognitive function"

–

*Supporting Information*

Marvin Petersen, MD; David Emskötter; Felix L. Nägele, MD; Carola Mayer, PhD; Maximilian Schell, MD; Jens Fiehler, MD; Jürgen Gallinat, MD; Simone Kuehn, PhD; Raphael Twerenbold, MD; Märit Jensen, MD; Götz Thomalla, MD; Bastian Cheng, MD

### Figure S1 – Sample selection flowchart

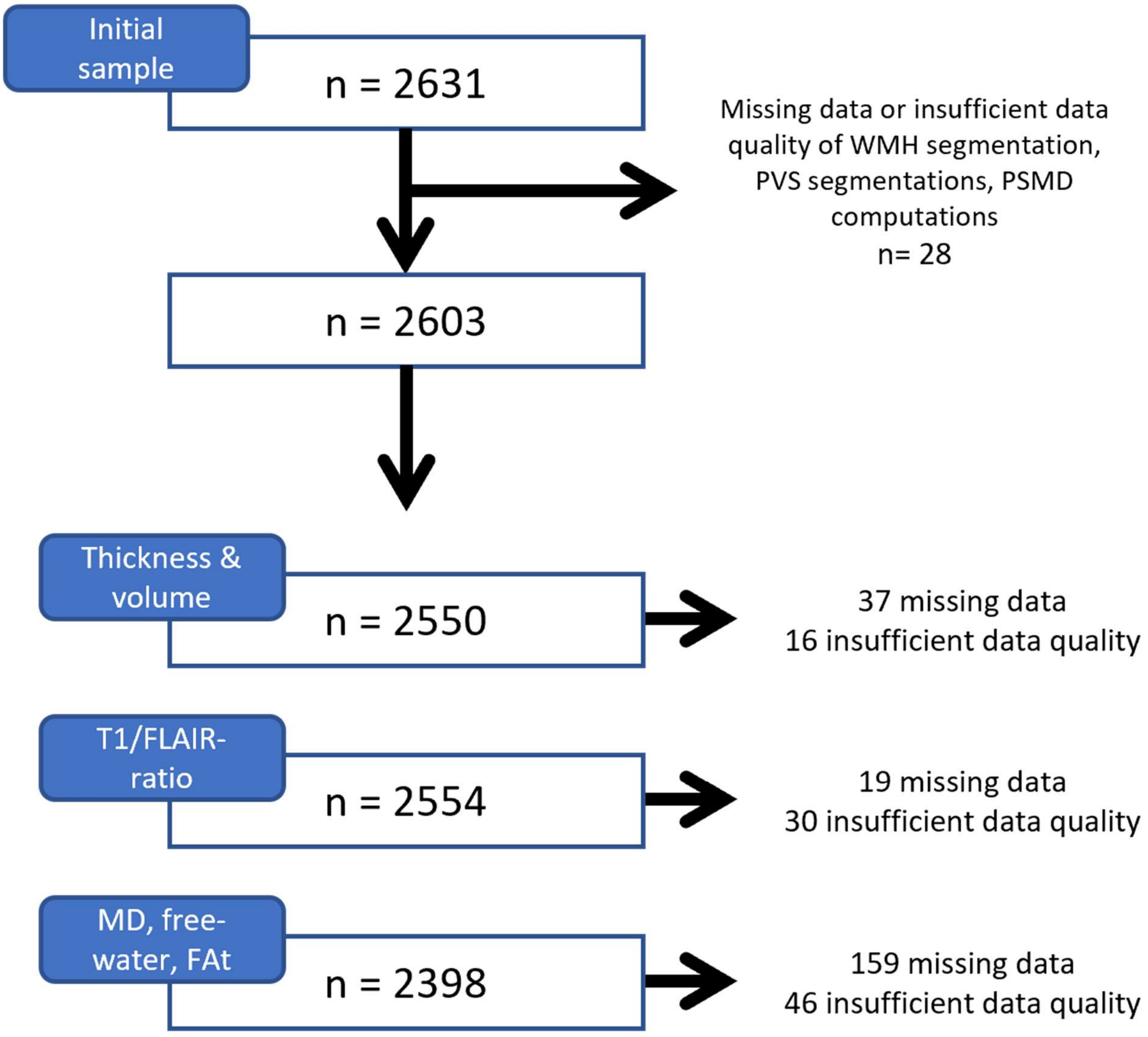

### Figure S2 – Correlation matrix of CSVD variables

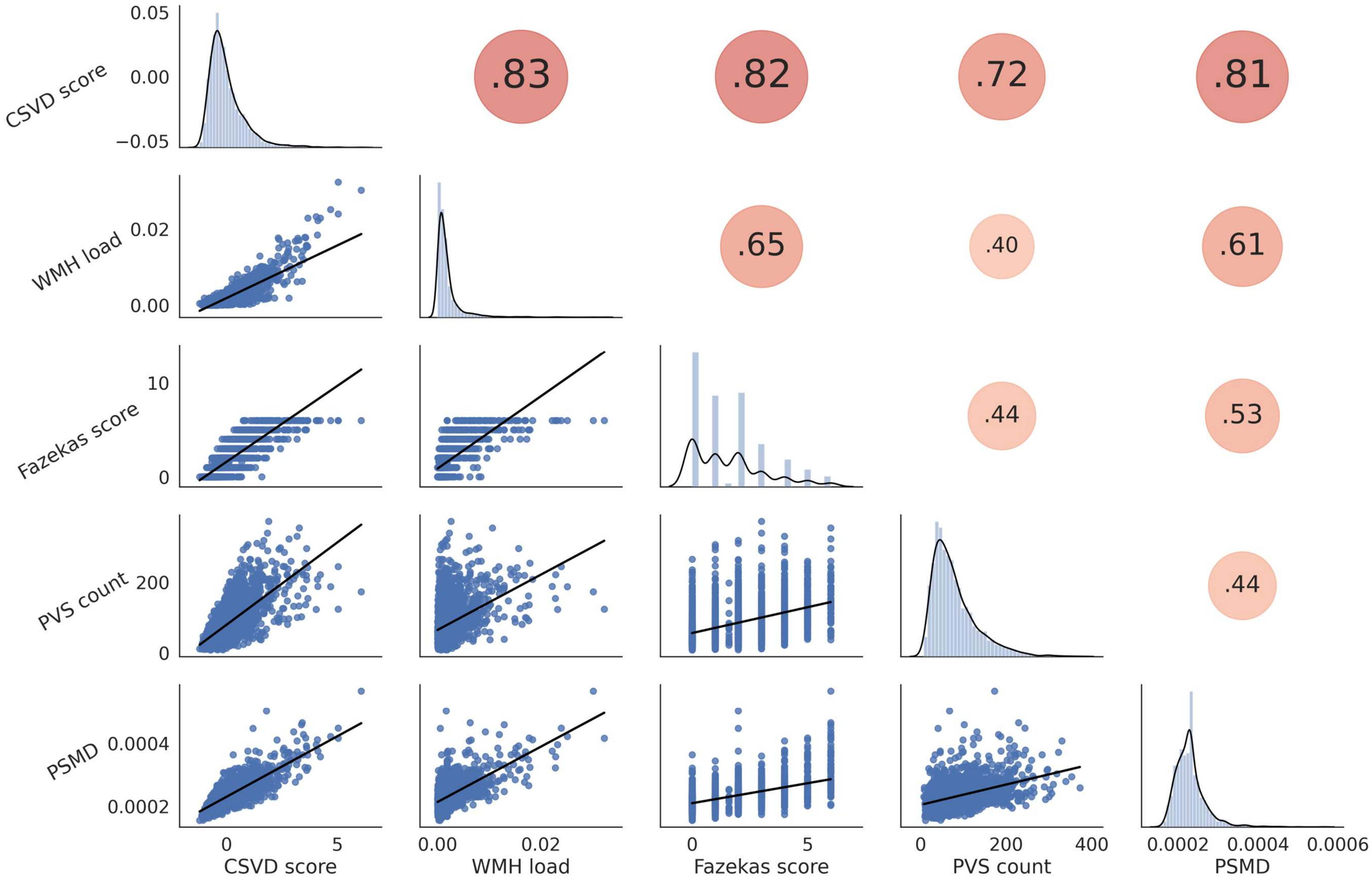

### Figure S3 – Correlation matrix of cognitive variables

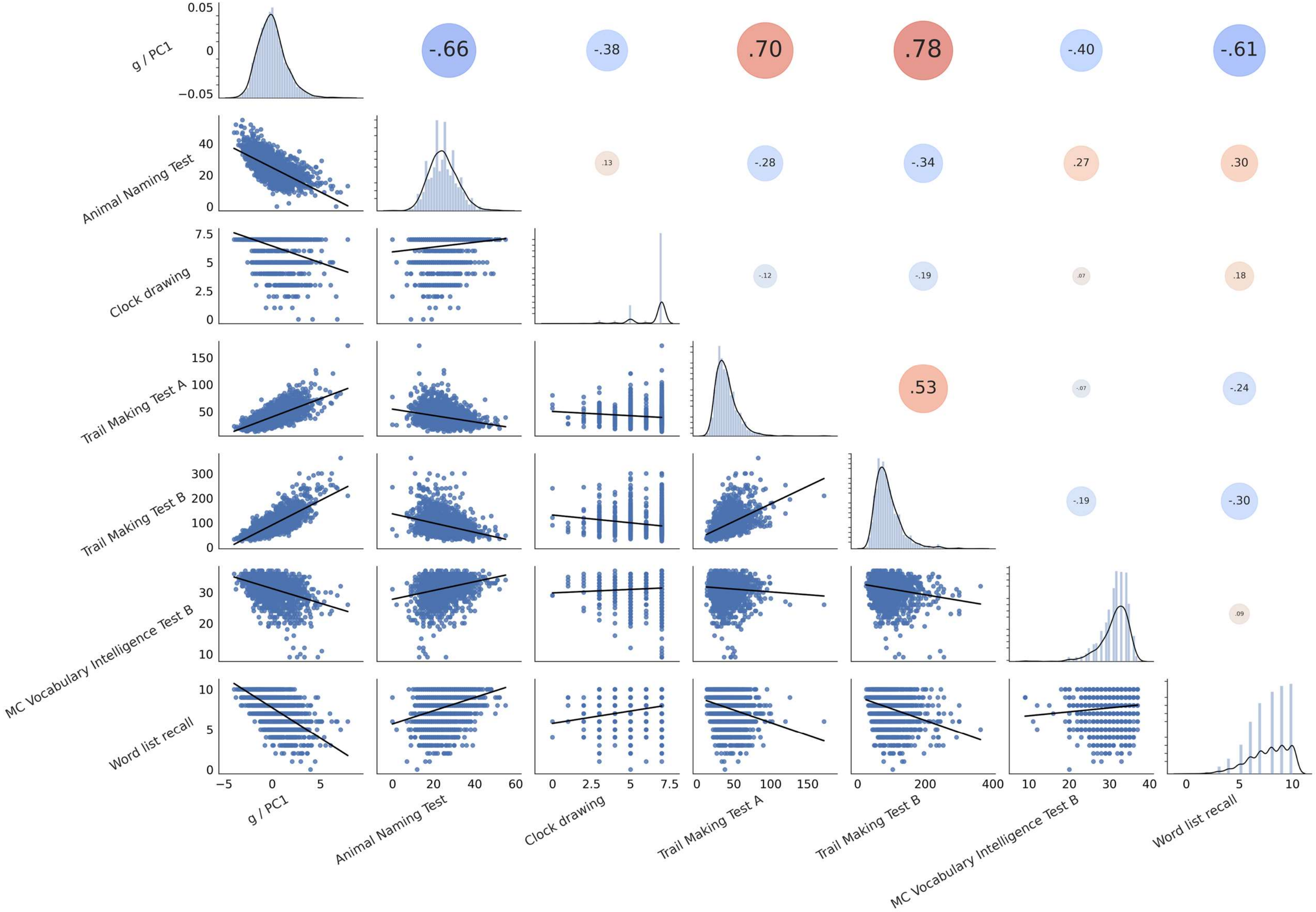

### Figure S4 – Cognition analysis individual cognitive test scores

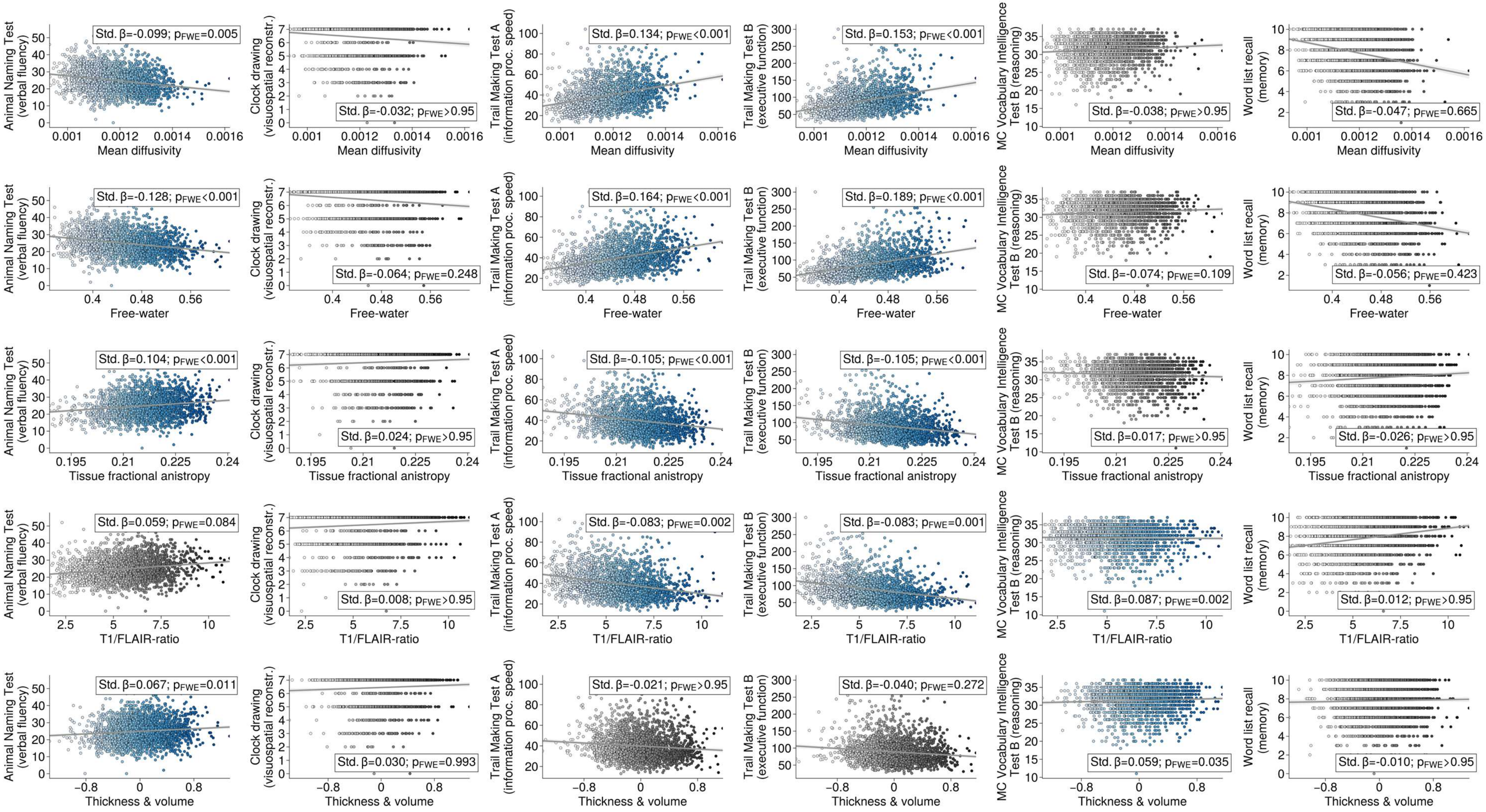

Regression plots displaying the association between aggregate imaging indices and cognitive test scores. For significant associations dots are highlighted in blue.
